# iSPAN: Improved prediction of outcomes post thrombectomy with Machine Learning

**DOI:** 10.1101/2023.04.17.23288611

**Authors:** Brendan S Kelly, Prateek Mathur, John Duignan, Sarah Power, Edward H Lee, Yuhao Huang, Silvia D Vaca, Laura M Prolo, Kristen W Yeom, Aonghus Lawlor, Ronan P Killeen

## Abstract

**Background:** This study aimed to develop and evaluate a machine learning model and a novel clinical score for predicting outcomes in stroke patients undergoing endovascular thrombectomy.

**Methods:** This retrospective study included all patients aged over 18 years with an anterior circulation stroke treated at a thrombectomy centre from 2010 to 2020. External validation data was obtained. The primary outcome variable was day 90 mRS ≥3. Existing clinical scores (SPAN and PRE) and Machine Learning (ML) models were compared. A novel clinical score (iSPAN) was derived by adding an optimised weighting of the most important ML features to the SPAN and compared results.

**Results:** 812 patients were initially included (397 female, average age 73), 62 for external validation. The best performing clinical score and ML model were SPAN and XGBoost (sensitivity specificity and accuracy 0.967, 0.290, 0.628 and 0.783, 0.693, 0.738 respectively). A significant difference was found overall and XGBoost was more accurate than SPAN (p< 0.0018). The most important features were Age, mTICI and total number of passes. The addition of 11 points for mTICI of ≤ 2B and ≥ 3 points for 3 passes to the SPAN achieved the best accuracy and was used to create the iSPAN. iSPAN was not significantly less accurate than XGBoost (p>0.5). In the external validation set, iSPAN and SPAN achieved sensitivity, specificity, and accuracy of (0.735, 0.862, 0.79) and (0.471, 0.897, 0.67), respectively.

**Conclusions:** iSPAN incorporates machine-derived features to achieve better predictions compared to existing scores. It is not inferior to the XGB model and is externally generalisable.

**Key Points:**

- An XGB model performed better than existing scores and other tested models for prognostication post EVT.
- It identified mTICI and number of passes as important and modifiable factors.
- Integrating these into the SPAN score (iSPAN) was not inferior to the XGB model and is generalisable and easier to use and interpret.

## Introduction

Acute ischaemic stroke is a time-dependent disease that continues to be a significant cause of morbidity and mortality worldwide ^1–3^. The advent of endovascular thrombectomy (EVT) has revolutionized acute stroke care, leading to improved patient outcomes ^4^. As a result, there has been a marked increase in the number of patients eligible for and undergoing EVT ^5^. However, a substantial proportion of patients do not achieve functional independence despite undergoing EVT. This makes the identification of patients who will have better outcomes post-EVT an important clinical challenge, both in terms of prospective patient selection and identification of patients at risk for poor outcome as soon as possible post thrombectomy.

To reach the agreed standard that at least 30% of patients should be functionally independent at 90 days post thrombectomy, as indicated by a 90-day modified Rankin score (mRS) of less than 3^6^, it is crucial to identify modifiable factors that may improve outcomes. Prognostication plays a vital role in guiding the selection of appropriate medical measures or surgical therapies for patients. Secondary data analysis of stroke databases has demonstrated the potential to identify practice-changing insights that could help improve patient selection for thrombectomy ^7^. Several potential prognostic factors have been suggested ^8,9^. While predictive scores have been proposed and trialled for this purpose ^10^, existing patient selection models are often based on data from patients who have already undergone EVT, resulting in selection bias and overly optimistic predictions of good outcomes. The integration of post-EVT information into selection scores may improve their prognostic utility, driving interest in the potential application of machine learning for predicting outcomes in these patients ^11^. However, preliminary studies in this area are limited by small sample sizes, with a recent systematic review of the topic including only 802 patients^12^. Further research with larger cohorts is necessary to ascertain the effectiveness of machine learning models in predicting outcomes for patients with stroke who undergo EVT.

Management of acute stroke has been disrupted by the advent of thrombolysis^13^, thrombectomy, and more recently by artificial intelligence in diagnostic^14^ and interventional radiology^15^. Despite the potential benefits of Artificial Intelligence (AI) in clinical settings, there are several barriers to its more widespread adoption ^16^. One notable challenge is the “black box” nature of many AI models, which can make it difficult for clinicians to understand and trust the decision-making process of these algorithms ^17^. This lack of transparency can hinder acceptance and limit the ability to identify potential sources of error ^18^. Additionally, integrating AI models into existing clinical software and workflows can be complex, requiring significant resources and technical expertise ^19^. Generalisability is also a concern, as AI models trained on specific patient populations or healthcare systems may not perform as well when applied to different contexts ^20^. Furthermore it has been shown that results of more complex stroke prediction models may not generalise to minority populations, and there have been recent calls for expansion of the pool of risk factors and improvements in modelling to mitigate against racial bias ^21^.

This study aimed to use machine learning to identify the variables that are predictive of an unfavourable outcome (90 day Modified Rankin Score (mRS) ≥3) in patients with an anterior circulation ischaemic stroke selected for EVT, and to determine if these features be combined with existing patient selection models to better predict outcome. Our hypotheses were that a machine learning model would identify patients post thrombectomy at higher risk of poor outcome and that a simpler model would be more generalisable.

## Methods

This retrospective cohort study received institutional review board approval and the need for prospective patient informed consent was waived under quality improvement grounds. This manuscript was prepared using the STROBE checklist^22^. All patients aged over 18 years with an anterior circulation stroke treated by our National Thrombectomy Service from 2010 to 2020 were included. All patients with an anterior circulation stroke treated at an international external tertiary referral centre between 2010 and 2018 were included for external validation. Both Pittsburgh Response to Endovascular therapy score and SPAN-100 score have been shown to be effective in similar studies and were used as performance comparison in this case ^23,24^.

### Statistical analysis and sample size calculation

The target variable was a binary prediction of mRS at 90 days extracted from the patients clinical chart (internal) or EPIC (external) and in all cases was calculated by a staff neurologist or stroke physician independently to the study. Sample size calculation was undertaken to develop a binary prediction model ^25^. A small optimism in predictor effect and estimates (as defined by a global shrinkage factor of ≥0.9) was used, as was with a small absolute difference of ≤0.05 in the model ‘s apparent and adjusted Nagelkerke ‘s R2. A precise estimation of the overall risk in the population was available. This showed that for a model with 4 predictive parameters a sample of at least 322 patients for development was required. Cochranes Q test was used to compare between multiple models and McNemar ‘s test was used in cases where two models were compared. Data were divided into an 80% 20% train test split. An external validation set was obtained to assess generalisability.

### Model training, evaluation and development of new predictive score

Several machine learning models (eXtreme Gradient Boosting (XGB), K-Nearest Neighbour (KNN), Support Vector Machine (SVM), Multilayer Perceptron (MLP)) were trained and tested and compared with two existing clinical scores (Stroke Prediction using Age and NIHSS (SPAN) and Pittsburgh Response to Endovascular therapy (PRE)). 10-fold cross validation was used to assess performance of the models in the training set.

Hyperparameter optimisation was employed using a grid search. The best performing optimized model was tested on a holdout set. For evaluation we computed and considered metrics including accuracy, sensitivity and specificity. Accuracy was used as the primary evaluation metric as our binary target class were relatively well balanced. Feature importance (gain based) were extracted using the plot_importance method in Scikit learn.

Feature importance was extracted from the best performing ML model and combined with best performing clinical score to create a novel *intelligence* clinical score. We then tested to determine non inferiority of the novel clinical score to the ML model. Finally the novel score was externally validated at an international institution and preliminary subgroup analysis was undertaken to assess for demographic effects.

## Results

A total of 812 patients were included in the initial analysis, with 397 being female and an average age of 73 (range 18-93) with 122 used for testing (62 with Day90-mRS of ≥3). For external validation, 63 patients were included, comprising 42 females and an average age of 74 (range 28-92) 29 with Day90-mRS of ≥3. Complete patient demographics are presented in Table 1. The best-performing clinical score and machine learning (ML) model were SPAN (sensitivity, specificity, accuracy) (0.967, 0.290, 0.628) and XGB (0.783, 0.693, 0.738). A significant difference in accuracy was observed overall, with XGB demonstrating significantly higher accuracy than the next best predictor, SPAN (p<0.0018). Comprehensive results can be found in Table 2.

**Table 1.**
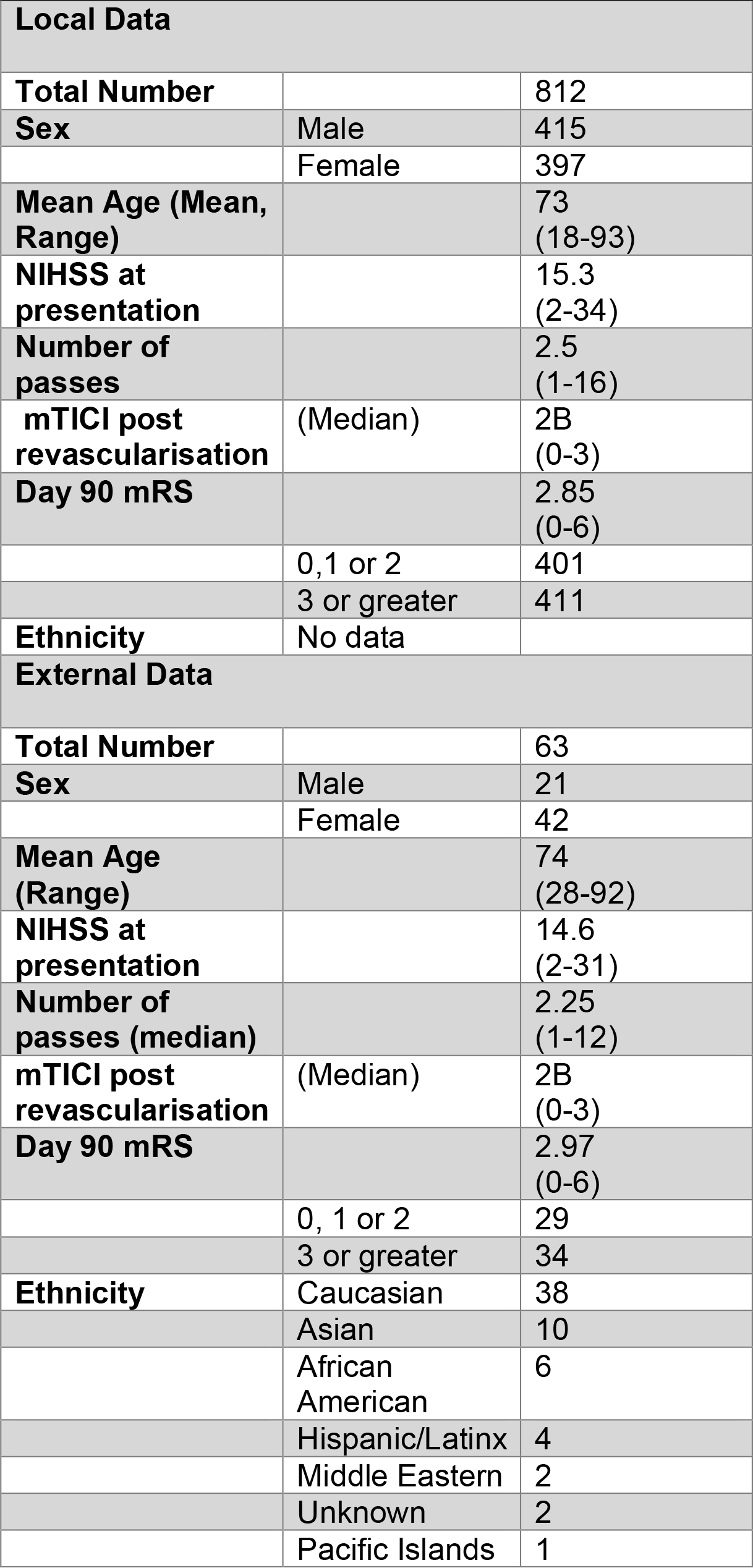
Patient demographic information

**Table 2.** Results of different models and clinical scores on test set

| Model | Accuracy | Sensitivity | Specificity |
| --- | --- | --- | --- |
| MLP | 0.612 | 0.533 | 0.689 |
| kNN | 0.566 | 0.59 | 0.541 |
| SVM | 0.545 | 0.933 | 0.163 |
| XGB | 0.738 | 0.783 | 0.693 |
| SPAN | 0.623 | 0.967 | 0.290 |
| iSPAN | 0.737 | 0.817 | 0.661 |

Feature importance of the XGB model is illustrated in Figure 1. SPAN can be calculated as (Age + NIHSS), with scores over 100 associated with poor outcomes. The most important features identified were Age, mTICI, and Total number of Passes. Given that mTICI and number of Passes are not included in SPAN, simulations were run to optimize the weight of their inclusion in the integrated SPAN (iSPAN) model (Figure 2). In the training set, adding 11 points for an mTICI of 2B or lower and a number of passes equal to or greater than 3 was found to be the optimal weighting for predicting poor outcomes. No significant difference in performance was observed between iSPAN and XGB on the internal test set (p>0.5) (Table 2).

**Figure 1.**
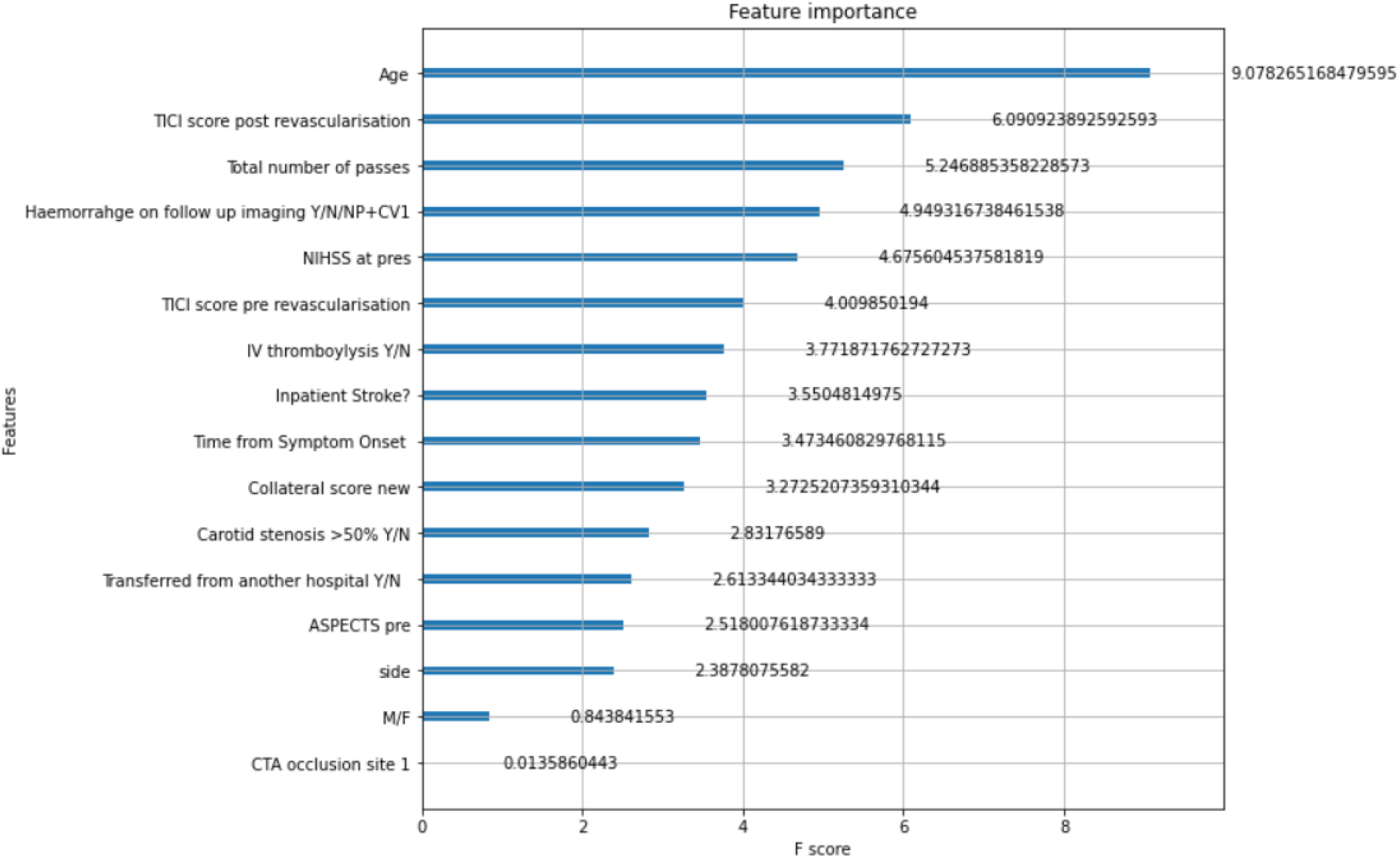
Title: XBG Feature Importance (Training) Legend: The feature importance with respect to information gain of the best preforming XGB model.

**Figure 2:**
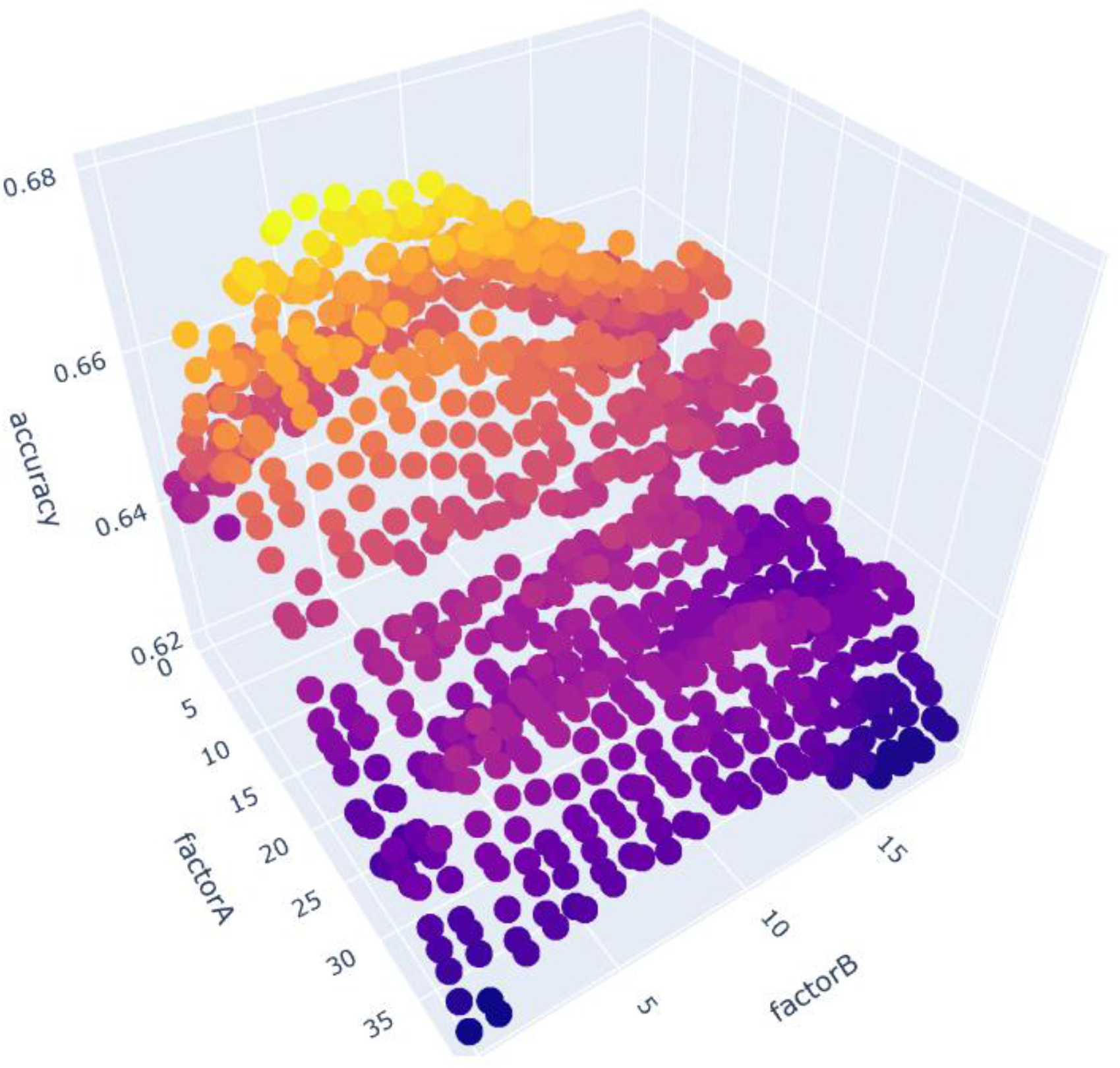
Title: Optimisation of iSPAN Legend: Training set weighting optimisation of mTICI (factorA) and number of passes (factorB)

In the external validation set, iSPAN and SPAN achieved sensitivity, specificity, and accuracy of (0.735, 0.862, 0.79) and (0.471, 0.897, 0.67), respectively. A comparison of the performance of SPAN and iSPAN on internal and external data is provided in Table 3. To help mitigate the propagation of racial or ethnic bias, the performance of iSPAN and SPAN on the external validation set for Caucasian patients compared to non-Caucasian patients is also presented.

**Table 3.** Results of iSPAN on test and validation sets

|  |  | SPAN | iSPAN |  |
| --- | --- | --- | --- | --- |
| **Internal** |  |  |  |  |
|  | Sensitivity | 0.967 | 0.817 |  |
|  | Specificity | 0.290 | 0.661 |  |
|  | Accuracy | 0.628 | 0.737 |  |
| **External** |  |  |  |  |
|  | Sensitivity | 0.897 | 0.735 |  |
|  | Specificity | 0.471 | 0.862 |  |
|  | Accuracy | 0.67 | 0.79 |  |
| **Sub Group** |  | SPAN | iSPAN | Relative change (%) |
| **Caucasian** |  |  |  |  |
|  | Sensitivity | 0.833 | 0.882 | 0.06 |
|  | Specificity | 0.654 | 0.81 | 0.29 |
|  | Accuracy | 0.71 | 0.84 | 0.18 |
| **Other** |  |  |  |  |
|  | Sensitivity | 0.40 | 0.66 | 0.4 |
|  | Specificity | 0.90 | 0.80 | -0.11 |
|  | Accuracy | 0.60 | 0.72 | 0.2 |

## Discussion

This study used an XBG model to identify features which can predict a poor outcome post thrombectomy. We also showed that integrating the features with the highest importance with an existing clinical score had similar performance to a more complicated ML model. The results demonstrate the potential utility of machine learning algorithms, such as XGB, and the integration of machine-derived additional variables in traditional clinical scores, in improving the accuracy and generalizability of outcome predictions for patients with stroke who undergo EVT. Our results are consistent with and build upon previous literature, which has shown the potential of machine learning algorithms to enhance prognostication in the context of acute ischemic stroke and EVT ^12^.

Predication of outcome post thrombectomy with machine learning is a common research question in the literature ^11,12,26^. Models such as XGB show state of the art performance on tabular data ^27^ and have been successful applied to smaller stoke databases ^28^. There is also hope that machine learning can add a level of personalisation to decision making around thrombectomy ^29^. However there is a large

### degree in heterogeneity in these studies ^12,29^

Many are also hampered by low sample size ^26,28,30–32^. Indeed a systematic review of the topic had a combined cohort of only 802 patients^12^, meaning that our internal cohort of 812 is a substantial contribution to the literature. Another advantage of our model is that we validated our results on an external population, unlike many other models that use internal or indeed no validation of their model ^26,28,30–32^. iSPANs performance metrics compare very favourably with published models especially when you consider its performance on external data.

The simplicity of iSPAN is another area where our model is different from the current literature. Even recent developments that focus strictly on interpretability suggest the use of additional algorithms to aid in explanations ^33^. We propose the addition of mTICI and the number of passes, which are both factors well known to influence outcome ^7,8,30,34^ and an intuitive reason for their effect on outcomes. That age, mTICI and number of passes a were the features identified as most important by machine learning also gives an intuition that our model is behaving as expected. Furthermore imaging factors such as ASPECTS and collaterals were weighted less highly which is concordant with the literature, however this is likely because of the inclusion criteria for thrombectomy.

The simplicity and interpretability of iSPAN offers key advantages over more complex machine learning models. Although a more complicated model might exhibit higher overall accuracy, a “black box” nature can impede clinicians ‘ and patients ‘ understanding and trust in the decision-making process^35^. In contrast, iSPAN integrates additional variables into the traditional SPAN score, making it more easily calculable, comprehensible and clinically applicable^36^. iSPAN ‘s ease of use and interpretability potentially renders it more appealing, as it allows for better insight into factors affecting a patient ‘s predicted outcome^35^. By providing a transparent scoring system, iSPAN promotes trust and confidence among healthcare professionals and patients and families, fostering its adoption in routine clinical practice^17,35,37^. Moreover, iSPAN ‘s simplicity (the addition of four numbers and comparing to 100) enables seamless integration into existing workflows, mitigating barriers often associated with complex AI models and legacy software systems^36^, crucial when “time is brain”. This effectively instant feedback is another advantage over other proposed models that require additional information over days or weeks ^28^.

There are two main avenues of clinical utility. First iSPAN can be used to give immediate prognostic information post thrombectomy to identify those patients at higher risk for a poor outcome. It is possible that such patients could benefit from more intense medical management. The second is the conformation of the importance of both maximising the mTICI while minimising the number of passes. Balancing these two factors is an important factor for interventional neuroradiologists to consider. It is also possible that this information prospectively could aid in selection of patients, for example where anatomical factors might make a higher number of passes more likely, but this would need prospective evaluation.

### Limitations and bias

There is a strong selection bias in our data as it includes only to patients who were selected for thrombectomy. As such it should only be applied to similar cohorts until further research shows prospective utility. No racial or ethnic data is available for the internal cohort but it is known that the overall population undergoing thrombectomy is overwhelmingly “White Irish” due to local population demographics. Performance on Caucasian compared to non-Caucasian patients is reported where available (on the external validation set) to help to mitigate against any further propagation of racial or ethnic bias. Many existing clinical stroke scores are known to perform worse on minority populations ^21^. While no statistically significant differences were found in performance between groups we were not powered to reject the null hypothesis. In absolute terms this seemed to be the case for SPAN in our study. However (again in absolute terms) our simple new iSPAN showed relatively better accuracy for non-Caucasian compared to Caucasian patients which suggests that our model at least does not further propagate any racial bias. We hypothesise that this is due to the universal and objective nature of the included features (mTICI and number of passes). This is in keeping with a recent large scale stroke prediction paper which concluded there is:

> *“(a) need to expand the pool of risk factors and improve modeling techniques to address observed racial disparities and improve model performance”* ^21^

## Conclusion

This study demonstrates the potential of both machine learning models, such as XGB, and the integration of additional variables into traditional clinical scores, like SPAN, to improve outcome predictions for patients with stroke undergoing EVT. The iSPAN model, in particular, offers a valuable alternative to “black box” machine learning models by combining enhanced predictive performance with simplicity, interpretability, and ease of use. These findings underscore the importance of refining prognostic tools to guide clinical decision-making, ultimately improving patient outcomes in acute ischemic stroke care.

## Data Availability

Data available on reasonable request

## Non standard abbreviations

(AI): Artificial Intelligence
(EVT): Endovascular thrombectomy
(XGB): eXtreme Gradient Boosting
(KNN): K-Nearest Neighbour
(SVM): Support Vector Machine
(ML): Machine Learning
(MLP): Multilayer Perceptron
(SPAN): Stroke Prediction using Age and NIHSS
(our funders): The added “I” in iSPAN refers to Improved, artificial Intelligence, Insight (our research centre), and the ICAT programme
(PRE): Pittsburgh Response to Endovascular therapy

